# VarPPUD: Variant post prioritization developed for undiagnosed genetic disorders

**DOI:** 10.1101/2024.04.15.24305876

**Authors:** Rui Yin, Alba Gutierrez, Undiagnosed Diseases Network, Shilpa Nadimpalli Kobren, Paul Avillach

## Abstract

Rare and ultra-rare genetic conditions are estimated to impact nearly 1 in 17 people worldwide, yet accurately pinpointing the diagnostic variants underlying each of these conditions remains a formidable challenge. Because comprehensive, *in vivo* functional assessment of all possible genetic variants is infeasible, clinicians instead consider *in silico* variant pathogenicity predictions to distinguish plausibly disease-causing from benign variants across the genome. However, in the most difficult undiagnosed cases, such as those accepted to the Undiagnosed Diseases Network (UDN), existing pathogenicity predictions cannot reliably discern true etiological variant(s) from other deleterious candidate variants that were prioritized through N-of-1 efforts. Pinpointing the disease-causing variant from a pool of plausible candidates remains a largely manual effort requiring extensive clinical workups, functional and experimental assays, and eventual identification of genotype- and phenotype-matched individuals. Here, we introduce VarPPUD, a tool trained on prioritized variants from UDN cases, that leverages gene-, amino acid-, and nucleotide-level features to discern pathogenic variants from other deleterious variants that are unlikely to be confirmed as disease relevant. VarPPUD achieves a cross-validated accuracy of 79.3% and precision of 77.5% on a held-out subset of uniquely challenging UDN cases, respectively representing an average 18.6% and 23.4% improvement over nine traditional pathogenicity prediction approaches on this task. We validate VarPPUD’s ability to discriminate likely from unlikely pathogenic variants on synthetic, GAN-generated candidate variants as well. Finally, we show how VarPPUD can be probed to evaluate each input feature’s importance and contribution toward prediction—an essential step toward understanding the distinct characteristics of newly-uncovered disease-causing variants.

**Significance Statement:** Patients with chronic, undiagnosed and underdiagnosed genetic conditions often endure expensive and excruciating years-long diagnostic odysseys without clear results. In many instances, clinical genome sequencing of patients and their family members fails to reveal known disease-causing variants, although compelling variants of uncertain significance are frequently encountered. Existing computational tools struggle to reliably differentiate truly disease-causing variants from other plausible candidate variants within these prioritized sets. Consequently, the confirmation of disease-causing variants often necessitates extensive experimental follow-up, including studies in model organisms and identification of other similarly presenting genotype-matched individuals, a process that can extend for several years. Here, we present VarPPUD, a tool trained specifically to distinguish likely from unlikely to be confirmed pathogenic variants that were prioritized across cases in the Undiagnosed Diseases Network. By evaluating the importance and impact of different input feature values on prediction, we gain deeper insights into the distinctive attributes of difficult-to-identify diagnostic variants. For patients who remain undiagnosed following comprehensive whole genome sequencing, our new method VarPPUD may reveal pathogenic variants amid a pool of candidate variants, thereby advancing diagnostic efforts where progress has otherwise stalled.

## Introduction

Rare and undiagnosed suspected genetic conditions cumulatively affect an estimated 25-30 million Americans and millions more worldwide.^1^ These diseases are typically associated with premature mortality or lifelong disability.^2,3^ However, each specific genetic condition may impact only a handful of individuals or as few as a single individual or family, hindering traditional case-control statistical approaches to study disease. As a result, diagnosing these conditions and uncovering disease-causing variants and mechanisms of action is extremely difficult. Indeed, patients suffering from rare disorders can spend multiple years and hundreds of thousands of dollars in pursuit of a diagnosis.^4^ Despite a steady increase in identifying the genetic aberrations that underlie rare Mendelian disorders, many conditions remain undiagnosed, in large part because patients’ presenting symptoms and disease-causing variants do not match known diseases.

To facilitate the diagnosis of rare and novel genetic disorders and further our collective understanding of the pathogenesis of these diseases, the United States’ National Institutes of Health funded the Undiagnosed Diseases Network (UDN) in 2014.^5–7^ Patients who are eventually referred to and accepted into the UDN undergo an extensive clinical evaluation with medical specialists at one of 12 clinical research sites across the country.^8^ The causes of undiagnosed disease are multifaceted, with genetic variants considered a primary factor.^9^ As such, in a majority of cases, UDN patients and their relevant affected and unaffected family members also receive whole genome sequencing. This data is computationally analyzed and filtered to select for variants that are rare, inherited in appropriate patterns, and have high functional impacts on phenotypically-relevant genes.^10^ Even after this process, not all prioritized variants are causal, and indeed most have no impact on health. Improved discernment of candidate variants that are eventually confirmed *pathogenic* is therefore of utmost importance.

Several computational tools have been developed to differentiate deleterious and pathogenic variants from benign variants in various disease contexts.^11–13^ These *in silico* approaches have utilized diverse features based on amino acid sequence and protein structure (e.g., PolyPhen2^14^), evolutionary conservation (e.g., MutationAssessor^15^, PROVEAN^16^), and phenotypic and other biomedical associations (e.g., SIFT^17^, MutationTaster^18^). Various underlying prediction models have also been explored, including hidden Markov models (e.g., FATHMM^19^), supervised machine learning (e.g., VEST^20^) and ensemble approaches (e.g., REVEL^21^, CADD^22^). These algorithms have been utilized extensively and successfully to prioritize disease-causing variants of uncertain significance (VUS) in clinical contexts, demonstrating the utility of predictive algorithms in diagnostic medicine.

Nevertheless, given a set of rare, functional, and clinically plausible candidate variants uncovered in an undiagnosed patient, existing pathogenicity predictors cannot easily distinguish disease-causing from non-causal or unlikely to be confirmed causal variants. Moreover, models underlying existing pathogenicity predictors tend to abstract away the contribution of each individual input feature when producing a final classification or score, making prediction interpretability and determination of variants’ functional impacts impossible.

Here, we present VarPPUD (**Var**iant **P**ost **P**rioritization for **U**ndiagnosed **D**isorders), a random forest-based model that classifies VUS as likely- or unlikely-to-be pathogenic based on gene-level functionality, nucleotide-level evolutionary constraint, physicochemical properties of amino acids, and existing variant deleteriousness predictions. VarPPUD is trained on a uniquely challenging set of real-world diagnostic and candidate variants that were uncovered in patients in the UDN through extensive manual efforts by clinical experts. Unlike other rare disease cohorts,^23^ the UDN accepts patients with both childhood- and adult-onset conditions that span a wide range of primary symptoms. A predictor trained on causal variants from this cohort would thus be generalizable across disease contexts. We show that VarPPUD is better able to discern likely pathogenic from other candidate variants relative to existing methods on this real-world dataset as well as on a synthetic dataset produced via a Generative Adversarial Network.^24–29^ Finally, VarPPUD’s underlying framework enables us to use Shapley Additive Explanation values^30^ to probe the contribution of each input feature and identify, for each prediction, the specific lines of evidence used to designate a candidate variant as likely pathogenic or not.

## Materials and Methods

### Variant compilation and pathogenicity classification

Upon acceptance to the UDN, each patient is assigned to one of 12 clinical sites where a team of medical, genetic, and bioinformatic specialists works collaboratively to find a diagnosis. These teams perform extensive in-person clinical evaluations of affected patients and relevant family members and analyze patients’ medical records, clinical reports, and prior or newly requested sequencing data to identify candidate disease-causing genetic variants.^10^ Clinical teams upload these prioritized genetic variants to the UDN Data Management and Coordinating Center and annotate them with a *status* of “solved”, “candidate”, or “rejected” and an *interpretation* of “pathogenic”, “likely pathogenic”, “uncertain significance”, “likely benign” or “benign” as recommended by the College of American Pathologists.^31^ All per-patient prioritized variants, standardized phenotype terms, demographic information and clinical data were subsequently loaded into a PIC-SURE instance for fast multi-modal querying (https://avillach-lab.hms.harvard.edu/pic-sure). Note that only confirmed pathogenic or highly likely pathogenic variants are submitted to ClinVar. From the 1733 patients evaluated in the UDN as of December 2021, we selected 474 SNV/indel variants with a status of “solved” or “candidate” and an available pathogenicity interpretation (Figure 1A). Each variant was prioritized in exactly one unique patient for a total of 474 patients considered. Population frequency analyses suggest that most variants annotated as “uncertain significance” are likely benign.^32^ Although some VUS may be disease causal, confirming their pathogenicity may be prohibitively difficult given a gene’s lack of model organism orthologs, patient’s exhibited phenotypes, and/or variant’s functional impact. We therefore reclassified all variants into two groups: 222 “likely pathogenic” variants with an original interpretation of “pathogenic” or “likely pathogenic”, and 252 “unlikely pathogenic” variants with an original interpretation of “uncertain significance”, “likely benign” or “benign”. Note that we use the phrase “unlikely pathogenic” to encompass both variants that are truly unlikely to be pathogenic as well as variants that are unlikely to be confirmed pathogenic in a short amount of time. Patient demographics grouped by prioritized variant category can be found in **Table 1**.

**Figure 1.**
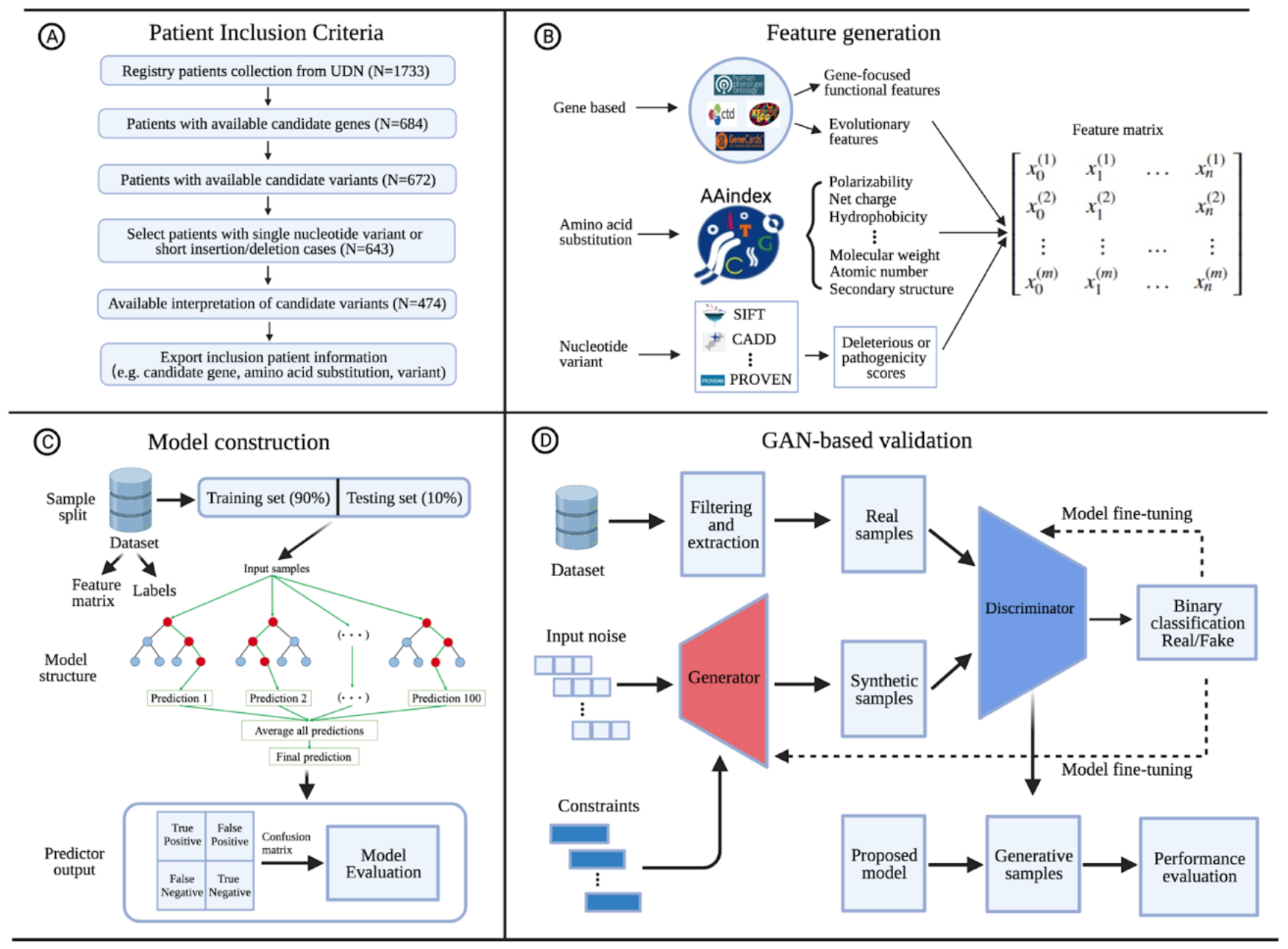
The workflow construction of the VarPPUD and its validation process for identifying strong pathogenic variants in UND patients. (**A) Procedure for patient inclusion in our study**. UDN patients lacking information about candidate genes, variants responsible for undiagnosed disorders, and unknown variant interpretations were excluded. Patients with long insertion/deletion (>5 nucleotides) or complicated variants on candidate genes were also filtered from the cohort. **(B)** Feature generation. We leveraged three categories i.e., gene-level functionality and evolutionary constraint, physicochemical properties of amino acids, and existing variant deleteriousness predictions, for feature generation. These features were acquired using various databases and tools based on the category of input data. The variables generated through distinct approaches were concatenated into a matrix for and fed into machine learning models. (**C) Model construction**. Filtered and processed samples were divided into training (90%) and testing (10%) datasets. Five-fold cross-validation was performed on the training set with a random forest classifier. The final performance estimated for the VarPPUD was quantified by metrics such as accuracy, precision, and recall, on the held-out testing test. **(D) GAN-based validation**. To validate the robustness of VarPPUD on potential new undiagnosed cases, we constructed a constraint-based generative adversarial network to produce synthetic testing data. We used the area under the Receiver Operator Characteristic (ROC) and Precision-Recall (PR) curves to evaluate the model’s performance.

### Gene-level features

We generated 12 gene-level features for each of the 474 variants in our real-world UDN dataset. These include: (1) number of unique phenotype terms associated with the gene from the Human Phenotype Ontology^33^ (HPO, https://hpo.jax.org/app/), (2) number of pathways that the gene appears in as listed in the Comparative Toxicogenomics Database^34^ (CTD, http://ctdbase.org/), (3) number of unique chemicals known to interact with the gene from CTD, (4) number of specific interactions between the gene and any chemical (i.e., chemical A inhibits the enzyme reaction of gene X, chemical A also reduces the expression of gene X) from CTD, (5) number of unique ways in which any chemical can interact with the gene (e.g., increases expression, affects folding, decreases reaction) from CTD, (6) number of rare diseases associated with the gene as listed in GeneAnalytics^35^ (https://geneanalytics.genecards.org/), (7) number of total diseases associated with the gene from GeneAnalytics, (8) evolutionary age of the gene from ProteinHistorian^36^ (https://proteinhistorian.docpollard.org/), (9) number of non-human species with an ortholog to the gene from phylogenetic profiles^37^ in ProteinHistorian, (10) dN/dS scores indicating selective direction and pressure from Evola^38^ (http://www.h-invitational.jp/evola/search.html), (11) essentiality scores indicating impact if both gene copies are lost from OGEE^39^ (https://v3.ogee.info/#/home), and (12) haploinsufficiency scores^40^ indicating impact if only one gene copy is lost.

### Amino acid-level features

We selected 21 continuous-valued physicochemical and biochemical properties defined for each amino acid type (e.g., polarity, molecular weight) from AAindex^41^ (https://www.genome.jp/aaindex/, Table S1). We determined the reference and alternate amino acid(s) for each prioritized variant wherever possible by retrieving their Human Genome Variation Society (HGVS) protein nomenclatures from LOVD (https://databases.lovd.nl/shared/variants) and MyGene2 (https://mygene2.org/MyGene2/genes).

For each amino acid property *x*, we compute an amino-acid based feature as Δ_*x*_*(ref, alt) = f*_*x*_*(alt) – f*_*x*_*(ref)*, where *f*_*x*_ is the value of property *x* for the query amino acid, *ref* is the reference amino acid sequence, and *alt* is the alternate amino acid sequence. We define *f*_*x*_*(stop) = 0* where *stop* is a termination codon and *f*_*x*_*(A,B*…,*Z) = f*_*x*_*(A) + f*_*x*_*(B) + … + f*_*x*_*(Z)* where *A, B*, and *Z* are individual amino acids. For frameshift variants, we set the reference amino acids to be the original protein sequence from the position of the first changed amino acid through the original termination codon, and the alternate amino acids to be the new sequence from the position of the first changed amino acid through the new termination codon. We did not encounter any variants that resulted in the loss of a termination codon, but for completeness for this case, we consider up to twenty amino acids past the original termination codon to be the alternate amino acids.

### Nucleotide-level features

For each prioritized variant, we queried its impacted gene, transcript, and annotated cDNA change in LOVD, MyGene2 and ClinVar of UDN at NIH (https://www.ncbi.nlm.nih.gov/clinvar/submitters/505999/) to determine the variant’s chromosome, genomic position, reference nucleotide(s) and alternate nucleotide(s) with respect to human genome build hg19/GRCh37. With variants in this format, we then computed deleteriousness or pathogenicity prediction scores for five other methods—SIFT^17^ (https://sift.bii.a-star.edu.sg/), VEST4^20^ (http://cravat.us/CRAVAT/), PROVEAN^16^ (http://provean.jcvi.org/index.php), FATHMM^19^ (http://fathmm.biocompute.org.uk/) and CADD^22^ (https://cadd.gs.washington.edu/)—using their default, recommended parameters. Note that the first two predictors score only missense variants, PROVEAN also scores in-frame indels, and the last two predictors additionally score frameshift indels, nonsense and non-coding splice-altering variants.

### Training the random forest classifier

We randomly split our 222 likely pathogenic and 252 unlikely pathogenic prioritized variants into training and validation sets in a 9:1 ratio, preserving the relative proportion of each variant type across the two sets. We then used Python’s Scikit-learn^42^ package to train a random forest classifier on the training set using 5-fold cross validation with bootstrapping to optimize model hyperparameters and reduce overfitting. Specifically, we randomly sampled with replacement from the training data to generate five folds—each with equivalent numbers of strongly and weakly pathogenic prioritized variants—and used each fold in turn as a held-out test set while training the model on the remaining four folds; we repeated this process 10 times. We also specified that each decision tree in the random forest model considered at most seven input features, minimized Gini impurity for each node split, had a maximum depth of five, had internal nodes with at least 30 samples, and had leaf nodes with at least 10 samples.

### Imputing missing feature values

Values for the 12 gene-based, 21 amino acid-based and 5 nucleotide-based features are missing in up to 37.6% of prioritized variants (Table 2). Because missing feature values may cause performance degradation in predictive models, we utilize Multivariate/Multiple Imputation in Chained Equations (MICE) to fill these in.^43,44^ Briefly, missing values for each feature are initialized with the mean of the non-missing values for that feature. Then, for each feature at a time, we fit a linear regressor using all other features as input (utilizing both the true observed and filled-in missing values) and the selected feature as output. We apply this fitted regressor to predict and fill in missing values for the selected feature. We then iteratively repeat this process over all features with missing values until convergence where values are no longer updated using the LinearRegression and IterativeImputer functions in Python’s Scikit-learn package. Imputing missing feature values boosted VarPPUD’s classification accuracy by 18.6%, precision (with respect to likely pathogenic variants) by 23.4%, and F-score by 14.5% during five-fold cross validation on the training set. Imputing missing values improved the performance of VarPPUD on the held-out validation set as well (Table 3). The final VarPPUD random forest model trained with imputed feature values had 340 individual decision trees.

**Table 1.**
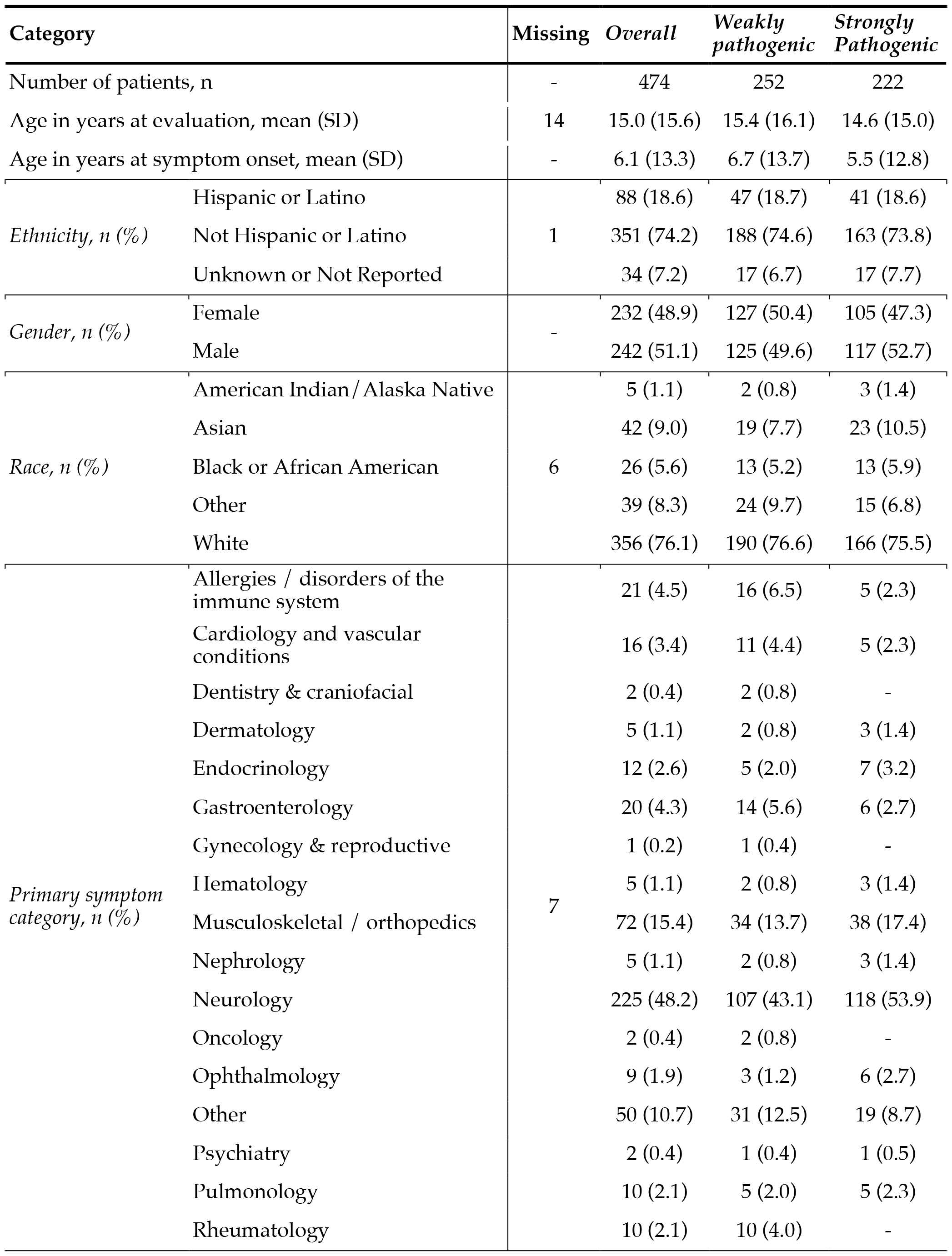
Demographic and clinical characteristics of patients grouped by the category of their prioritized variant. Prioritized genetic variants are assigned to either the “weakly pathogenic” or “strongly pathogenic” categories based on their provided clinical interpretation. Each prioritized variant corresponds to a single affected patient.

**Table 2.** Feature value properties for UDN-prioritized variants. Ranges for integer feature values are denoted by “#” and listed with a “..” separator between minimum and maximum allowed values, and ranges for float feature values are listed with a “,” separator instead. Percent missing values are calculated out of all 474 prioritized variants. Amino acid-based features are computed as the change in values between reference and alternate alleles; see Methods for further details.

| Type | Feature | Range | Missing | Mean | Standard Deviation |
| --- | --- | --- | --- | --- | --- |
| *gene-based* | # phenotypes associated with gene | {0 .. +∞} | 0 (0%) | 172 | 169 |
|  | # pathways gene is part of | {0 .. +∞} | 0 (0%) | 13.7 | 25.8 |
|  | # unique chemicals gene interacts with | {0 .. +∞} | 0 (0%) | 36.4 | 34.2 |
|  | # total chemical-gene interactions | {0 .. +∞} | 0 (0%) | 47.1 | 66.9 |
|  | # chemical-gene interaction types | {0 .. +∞} | 0 (0%) | 11.4 | 10.6 |
|  | # rare diseases associated with gene | {0 .. +∞} | 0 (0%) | 21.3 | 24.4 |
|  | # total diseases associated with gene | {0 .. +∞} | 0 (0%) | 30 | 36.9 |
|  | evolutionary age | [0, +∞) | 42 (8.9%) | 989 | 837 |
|  | # species with an ortholog | {0 .. +∞} | 42 (8.9%) | 2.51 | 3.07 |
|  | dN/dS | [0, 1] | 68 (14.3%) | 0.18 | 0.162 |
|  | gene essentiality | {0 .. 2} | 45 (9.5%) | 0.455 | 0.543 |
|  | gene haploinsufficiency | [0, 1] | 142 (30.0%) | 0.496 | 0.251 |
| *amino acid-based* | polarity | [-2.54, 2.54] | 32 (6.8%) | -0.261 | 0.948 |
|  | net charge | {-2 .. 2} | 32 (6.8%) | 0.0566 | 0.712 |
|  | hydrophobicity | [-3.37, 3.37] | 32 (6.8%) | -0.181 | 1.16 |
|  | Van der Waals volume | [-8.08, 8.08] | 32 (6.8%) | -0.185 | 2.54 |
|  | polarizability | [-0.409, 0.409] | 32 (6.8%) | -0.0112 | 0.122 |
|  | pK-COOH | [-1.2, 1.2] | 32 (6.8%) | -0.245 | 0.721 |
|  | pK-NH2 | [-2.29, 2.29] | 32 (6.8%) | -0.526 | 2.32 |
|  | hydration | [-5.2, 5.2] | 32 (6.8%) | -0.266 | 1.68 |
|  | molecular weight | [-129.17, 129.17] | 32 (6.8%) | -7.41 | 49.4 |
|  | optical rotation | [-100.8, 100.8] | 32 (6.8%) | 5.11 | 31.3 |
|  | secondary structure | {-2 .. 2} | 32 (6.8%) | -0.219 | 1.13 |
|  | free energy solution | [-7.15, 7.15] | 32 (6.8%) | -0.284 | 2.08 |
|  | # hydrogen bonds | {-4 .. 4} | 32 (6.8%) | 0.299 | 1.78 |
|  | residue volume | [-169.7, 169.7] | 32 (6.8%) | -6.06 | 63.8 |
|  | transfer free energy | [-2.7, 2.7] | 32 (6.8%) | -0.203 | 0.929 |
|  | side chain interaction | [-5.64, 5.64] | 32 (6.8%) | -0.622 | 2.36 |
|  | # vertices | {-10 .. 10} | 32 (6.8%) | -0.233 | 3.11 |
|  | # edges | {-12 .. 12} | 32 (6.8%) | -0.258 | 3.36 |
|  | eccentricity | [-11.1, 11.1] | 32 (6.8%) | -0.221 | 3.52 |
|  | diameter | [-14, 14] | 32 (6.8%) | -0.0475 | 5.02 |
|  | atomic number | [-61, 61] | 32 (6.8%) | -2.64 | 18 |
| *nucleotide-based* | SIFT | [0, 1] | 42 (8.9%) | 0.154 | 0.311 |
|  | VEST | [0, 1] | 129 (27.2%) | 0.671 | 0.291 |
|  | PROVEAN | (-∞, +∞) | 178 (37.6%) | -3.39 | 2.68 |
|  | FATHMM | [0, 1] | 67 (14.1%) | 0.696 | 0.38 |
|  | CADD | (-∞, +∞) | 42 (8.9%) | 3.17 | 2.18 |

**Table 3.**
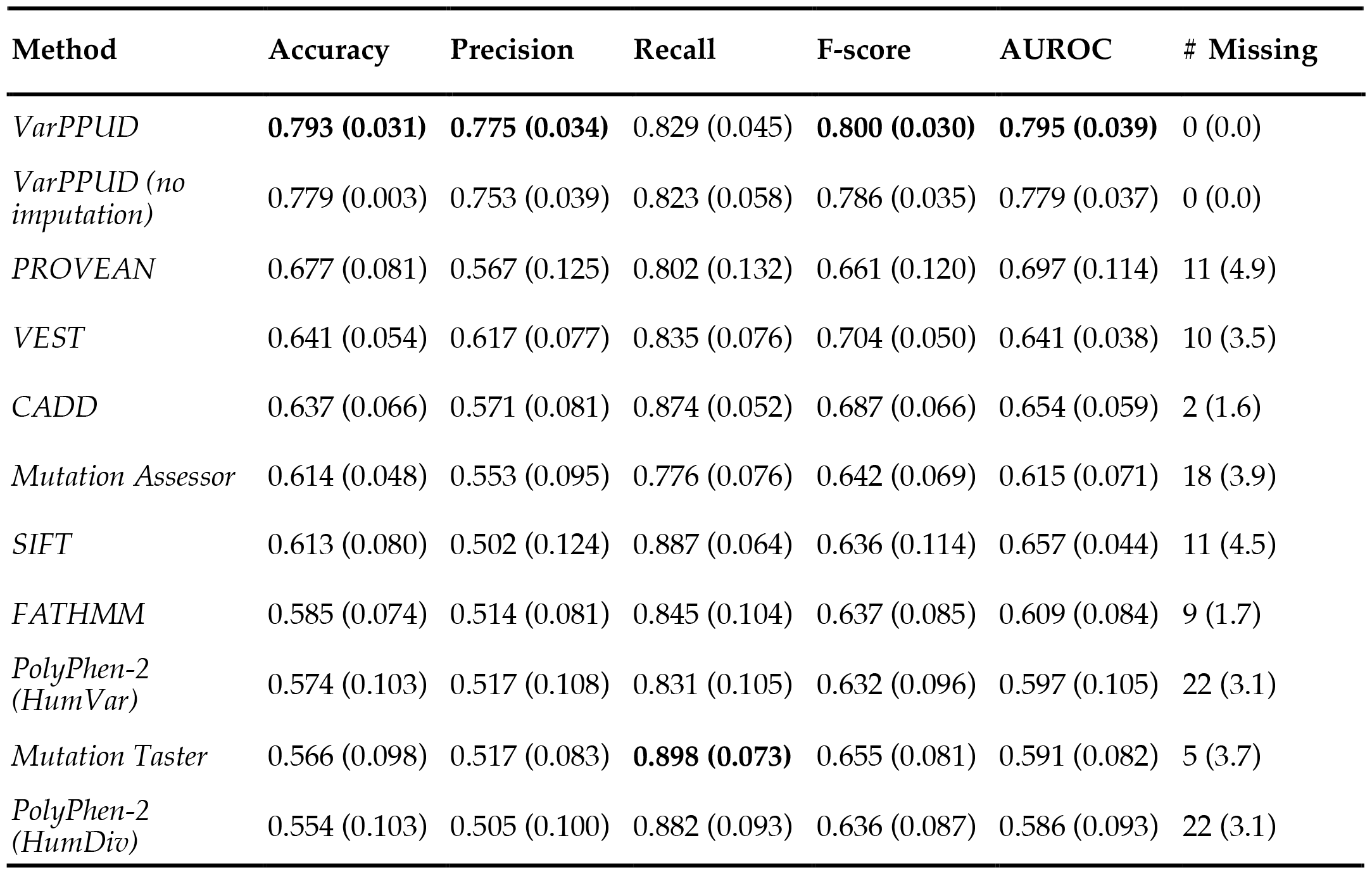
Performance in distinguishing strongly (positive) from weakly (negative) pathogenic variants in real-world validation set. Performance comparison between two versions of VarPPUD (i.e., with and without feature value imputation) and previous methods for distinguishing strongly from weakly pathogenic variants in real-world patients from the UDN. The number of “missing” variants indicates the number of variants in the validation test that the given method could not generate a prediction for.

| Method | Accuracy | Precision | Recall | F-score | AUROC | # Missing |
| --- | --- | --- | --- | --- | --- | --- |
| *VarPPUD* | **0.793 (0.031)** | **0.775 (0.034)** | 0.829 (0.045) | **0.800 (0.030)** | **0.795 (0.039)** | 0 (0.0) |
| *VarPPUD (no imputation)* | 0.779 (0.003) | 0.753 (0.039) | 0.823 (0.058) | 0.786 (0.035) | 0.779 (0.037) | 0 (0.0) |
| *PROVEAN* | 0.677 (0.081) | 0.567 (0.125) | 0.802 (0.132) | 0.661 (0.120) | 0.697 (0.114) | 11 (4.9) |
| *VEST* | 0.641 (0.054) | 0.617 (0.077) | 0.835 (0.076) | 0.704 (0.050) | 0.641 (0.038) | 10 (3.5) |
| *CADD* | 0.637 (0.066) | 0.571 (0.081) | 0.874 (0.052) | 0.687 (0.066) | 0.654 (0.059) | 2 (1.6) |
| *Mutation Assessor* | 0.614 (0.048) | 0.553 (0.095) | 0.776 (0.076) | 0.642 (0.069) | 0.615 (0.071) | 18 (3.9) |
| *SIFT* | 0.613 (0.080) | 0.502 (0.124) | 0.887 (0.064) | 0.636 (0.114) | 0.657 (0.044) | 11 (4.5) |
| *FATHMM* | 0.585 (0.074) | 0.514 (0.081) | 0.845 (0.104) | 0.637 (0.085) | 0.609 (0.084) | 9 (1.7) |
| *PolyPhen-2 (HumVar)* | 0.574 (0.103) | 0.517 (0.108) | 0.831 (0.105) | 0.632 (0.096) | 0.597 (0.105) | 22 (3.1) |
| *Mutation Taster* | 0.566 (0.098) | 0.517 (0.083) | **0.898 (0.073)** | 0.655 (0.081) | 0.591 (0.082) | 5 (3.7) |
| *PolyPhen-2 (HumDiv)* | 0.554 (0.103) | 0.505 (0.100) | 0.882 (0.093) | 0.636 (0.087) | 0.586 (0.093) | 22 (3.1) |

### Generating synthetic testing data

We utilized the generative adversarial network-based method CTGAN^28^ to generate additional synthetic variants from the unimputed feature vectors of UDN prioritized variants. We specified that all feature values for synthesized data must be within their appropriate ranges and of the correct data type (i.e., integer or float, Table 2) with reasonable constraint design due to the correlation between features. For example, the number of associated rare diseases cannot be greater than the number of total associated diseases. Just like the real-world data they were based on, the resulting 10,000 synthetic variants had some missing feature values, so we randomly selected 250 likely pathogenic and 250 unlikely pathogenic synthetic variants with no missing feature values. We then randomly divided these variants into five groups with 50 likely pathogenic and 50 unlikely pathogenic synthetic variants each, and evaluated VarPPUD’s predictive performance on each of these five synthetic test sets.

### Testing alternate pathogenicity predictors

We compared VarPPUD to nine related variant pathogenicity predictors, five of which were used as input features in our model (i.e., SIFT, VEST4, PROVEAN, FATHMM, and CADD). We also retrieved variant pathogenicity predictions from PolyPhen-2 (HumDiv and HumVar)^14^ (http://genetics.bwh.harvard.edu/pph2/bgi.shtml), Mutation Taster^45^ (https://www.mutationtaster.org/) and Mutation Assessor^15^ (http://mutationassessor.org/r3/). We considered likely and unlikely pathogenic variants to be positive and negative examples respectively to compute predictive accuracy, precision, recall, F1 score, and area under the receiver operating characteristic curve (AUROC) for each method on the held-out validation set. We also evaluated our model through area under the precision–recall curve (AUPRC) for the external validation with synthetic dataset.

### Measuring feature contribution

We measure how each of our 38 input features contributes toward prediction on our real-world validation set in two ways. First, we randomly shuffled the values for each feature in turn and measured how much overall accuracy on the validation set dropped using the permutation_importance function from Python’s Scikit-learn package. Then, we utilized the Shapley Additive exPlanations (SHAP) package to compute and visualize Shapley values for each individual prediction.^30,46^ Negative Shapley values indicate a feature’s contribution toward predicting a variant to be unlikely pathogenic, whereas positive Shapley values indicate a feature’s contribution toward predicting a variant to be likely pathogenic. To determine the overall (rather than per-prediction) contribution of each feature, we computed the mean of the absolute values of each feature’s SHAP values across all variants in the validation set. Finally, we computed SHAP interaction values, capturing how pairs of feature values contribute to prediction.

### Incorporating patient demographic features

Each of the starting 222 likely pathogenic and 252 unlikely pathogenic variants from real-world UDN patients are associated with additional patient information, including age at first symptom onset, “current” age at evaluation, self-identified gender, self-identified ethnicity (Hispanic/Latino or otherwise), and self-identified race. For each of these features, we retrained VarPPUD ten times using the original 38 features and the new demographic feature. Each retraining used a different random seed to split the starting variants into training and validation sets in a 9:1 ratio. Changes to predictive performance was measured as the change in area under the receiver-operator curve compared to the version of VarPPUD with no demographic features.

## Results

### Patient demographics

We trained a new variant pathogenicity predictor, VarPPUD, on 474 unique variants impacting 413 genes that were originally prioritized through extensive manual and computational efforts in 474 patients enrolled in the Undiagnosed Diseases Network (UDN).^10^ These patients’ symptoms began at age 6.1 on average, and they endured multiple failed diagnostic attempts for nearly 9 years on average before being accepted into and evaluated by the UDN (Table 1). As expected, the genetic variants eventually prioritized in their cases are extremely unique and often had never been previously identified as disease-causing.^9^ The candidate and solved genetic variants from these patients were designated by clinical teams as likely pathogenic (n=222) or otherwise (called unlikely pathogenic [n=252] herein); VarPPUD was trained to discriminate between these two classes (see Methods for details). Our cohort of patients included an equivalent number of female and male patients (232 [48.9%] vs. 242 [51.1%]) with a majority classified as white (356 [76.1%]). Patients’ primary symptoms in both the likely and unlikely pathogenic sets were mostly neurologic (118 [53.9%] vs. 107 [43.1%], respectively) or musculoskeletal (38 [17.4%] vs. 34 [13.7%], respectively). Variants in patients with primarily rheumatologic (10), oncologic (2) or craniofacial (2) symptoms were exclusively in the unlikely pathogenic class. Overall, patients’ primary symptom categories between the two variant classes were evenly distributed, with an average ratio of 1:1.2 between the likely and unlikely pathogenic variant classes across all primary symptom categories.

### VarPPUD’s predictive ability on real-world and synthetic variants

We held out 10% of the real-world prioritized variants as a validation set, and used the remaining 90% of the data to train and test VarPPUD. Our model achieved an overall classification accuracy of 0.793 and a precision in detecting likely pathogenic variants of 0.775 on the validation set (Table 3). Due to the small number of variants in our validation set, we also generated five sets of synthetic variants using a GAN-based technique (see Methods for details); each synthetic test set contained 50 likely and 50 unlikely pathogenic variants. VarPPUD achieved an area under the receiver-operator characteristic curve (AUROC) of 0.826 to 0.891 (0.860 on average) across these synthetic test sets, and an area under the precision-recall curve (AUPRC) of 0.807 to 0.887 (0.848 on average) with respect to likely pathogenic variants (Figure 2).

**Figure 2.**
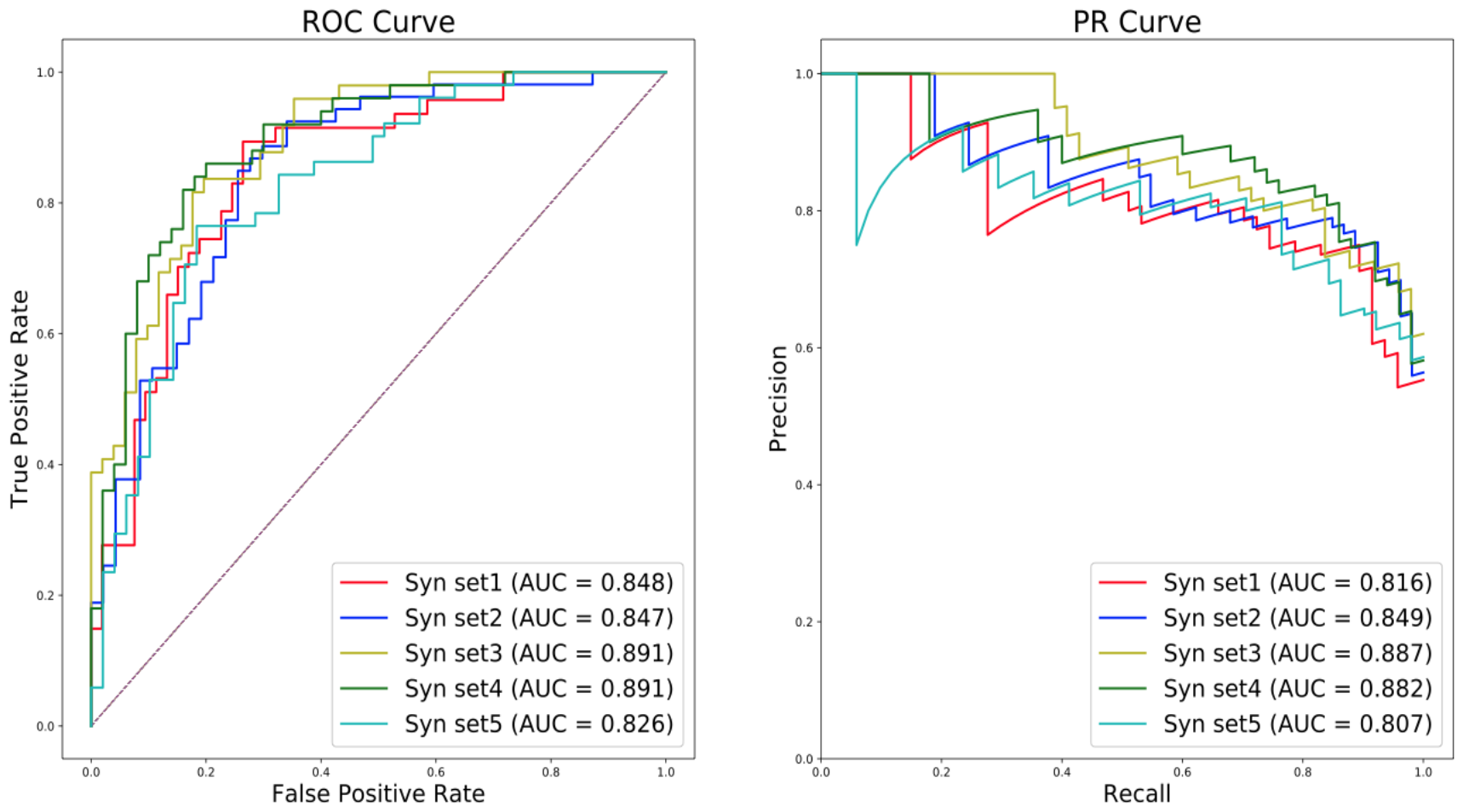
VarPPUD’s predictive performance on five GAN-synthesized variant datasets. Performance of the proposed model at identifying strong pathogenic variants on five different GAN-based synthetic testing sets evaluated by Receiver Operating Characteristic (ROC) and Precision-Recall (PR) curves.

These results suggest that VarPPUD can stably discriminate between likely and unlikely pathogenic variants in both real-world and synthetic datasets.

### Comparison to related pathogenicity predictors

VarPPUD is trained on a unique set of variants that were all initially prioritized due to their potential to cause disease; discriminating between likely and unlikely pathogenic variants within this set is a goal that existing pathogenicity predictors were not specifically designed nor optimized for. Moreover, some existing pathogenicity predictors only generate predictions for specific variant types (e.g., PolyPhen-2, SIFT), and therefore could not generate predictions for two (CADD) to 22 (PolyPhen-2) variants in our validation set. Nevertheless, we sought to compare VarPPUD’s ability to distinguish likely from unlikely pathogenic variants across all variant types—including large protein-coding indels, nonsense and synonymous variants—to the ability of nine related pathogenicity predictors to perform this same task. Estimating the pathogenicity of all variant types is especially desirable in still-undiagnosed cases where known, disease-causing, protein-altering SNV/indel variants were not uncovered. To this end, we computed pathogenicity or deleteriousness predictions for all variants in our validation set, where possible, using SIFT,^17^ VEST4,^20^ PROVEAN,^16^ FATHMM,^19^ CADD,^22^ Mutation Taster,^45^ Mutation Assessor,^15^ and two versions of PolyPhen-2 (i.e., disease variants are differentiated from variants at divergent sites across close mammalian homologs [HumDiv] or from common human polymorphisms unassociated with disease [HumVar]).^14^ We found that VarPPUD achieved higher overall prediction accuracy, higher precision in identifying likely pathogenic variants, a higher F-score, and a higher AUROC than all other methods on the held-out validation set of real-world prioritized variants (Table 3). However, all but two methods (PROVEAN and Mutation Assessor) achieved higher recall of likely pathogenic variants than VarPPUD, with Mutation Taster achieving the highest recall across all methods. As previously mentioned, these alternate predictors were trained to distinguish pathogenic from *benign* (rather than from unlikely pathogenic) variants across the whole genome, rather than within a prioritized variant list, and therefore would predict nearly all the variants in our validation set to be likely pathogenic. As such, it is expected that they would have higher recall of likely pathogenic variants within this set.

Nevertheless, VarPPUD achieved a comparably high recall of 0.829 on our validation set. Moreover, we found that although the imputation of missing feature values improved VarPPUD’s predictive performance, even the version of VarPPUD without feature value imputation better identified likely pathogenic variants relative to other existing tools on nearly all evaluation metrics. These results demonstrate the relatively superior ability of VarPPUD to distinguish likely from unlikely pathogenic variants within a set of prioritized, candidate disease-causing variants.

### Importance and contribution of VarPPUD features

We next wanted to evaluate the importance of each of VarPPUD’s features. VarPPUD incorporates 12 gene-level features (e.g., number of associated diseases or pathways, evolutionary age, essentiality and haploinsufficiency), 21 features capturing putative physicochemical changes due to amino acid substitutions, and five deleteriousness scores from existing *in silico* methods (see Methods for details). Although some amino acid-based features were highly correlated with each other as expected (e.g., polarity decreases as number of hydrogen bonds increases, Pearson Correlation Coefficient [PCC] > 0.9, Figure 3a), most features showed low correlations with each other, particularly between features from different categories (−0.1 < PCC < 0.1). The five *in silico* deleteriousness features tended to be most highly correlated with each other, with CADD, FATHMM, and VEST4 showing the lowest relative correlations amongst each other and SIFT and PROVEAN showing the highest relative correlations. Overall, most of VarPPUD’s input features are uncorrelated and thus have the potential to contribute complementary information relevant for variant pathogenicity prediction.

**Figure 3.**
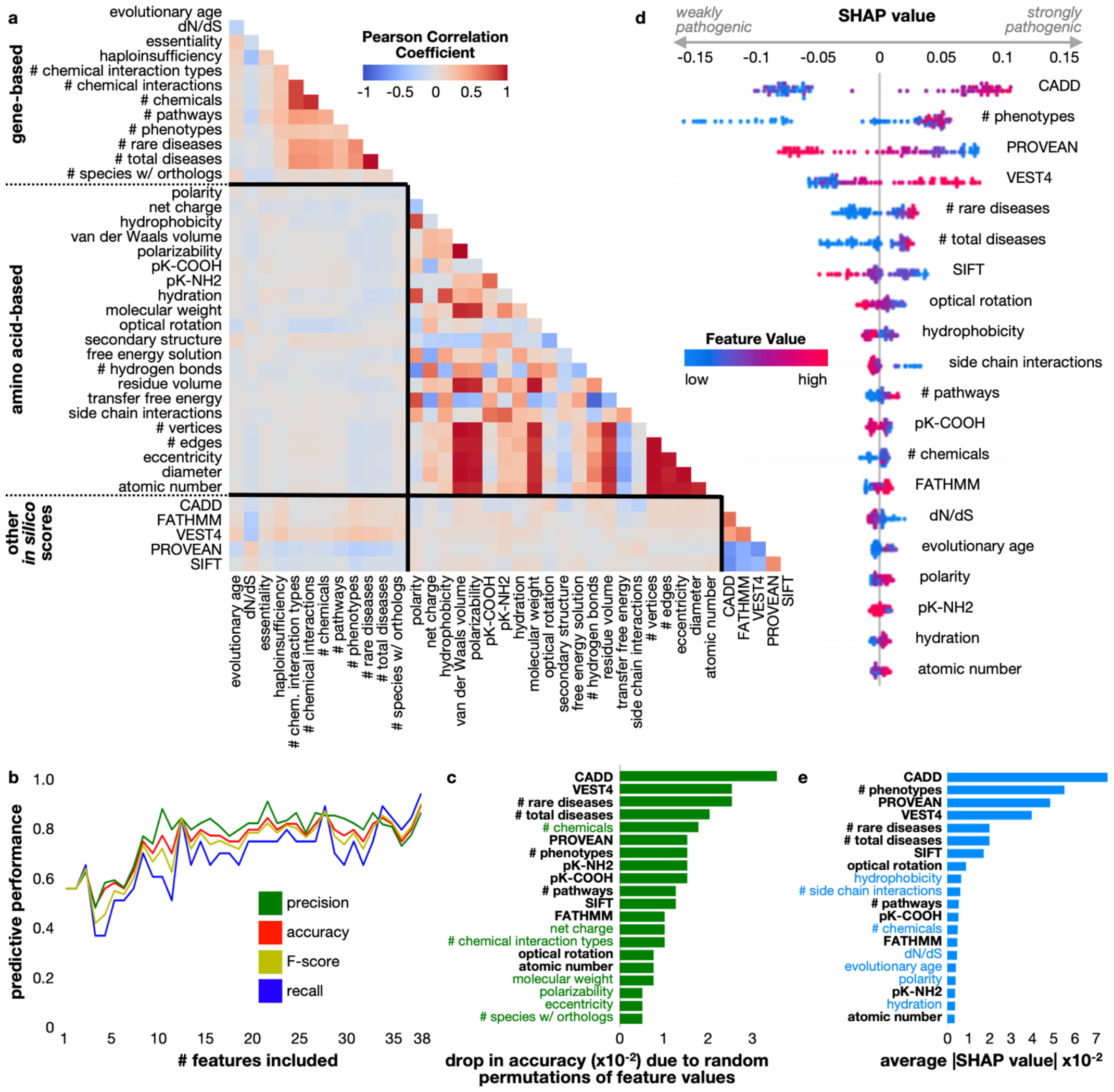
Feature correlation and contribution toward prediction. **(a)** Correlations between feature values were computed for all variants in the real-world training dataset using Pearson’s Correlation Coefficient. Feature names are grouped by category: gene-based, amino acid-based, and nucleotide-based (*in silico* deleteriousness predictions) and separated by black lines in the heatplot. **(b)** Features were eliminated in the same order as Table 2 and the model was retrained with five-fold cross validation on the training set; accuracy (red), precision (green), recall (blue), and F-score (yellow) were computed from performance on the held-out test set. **(c)** Feature importance measured as the drop in accuracy on the test set when values for that feature were randomly permuted in the training set. Feature names highlighted in green indicate importance by this measure and not by the measure in (e). **(d)** SHAP values for each feature in each individual prediction on the real-world test set. Negative SHAP values indicate contribution toward predicting a variant to be weakly pathogenic, whereas positive SHAP values indicate a feature’s contribution toward predicting a variant to be strongly pathogenic. Points are colored by their linearly normalized (between 0 and 1) feature value, organized from low (blue) to high (red). **(e)** Feature importance measured as the average of the absolute values of the SHAP values computed for all variants in the test set; feature names highlighted in blue indicate importance by this measure and not be the measure described in (d).

We first investigated how individual features contribute to VarPPUD’s overall discriminatory power. To this end, we trained ablated versions of VarPPUD using subsets of input features and evaluated the resulting models’ performance using precision, accuracy, F-score and recall of likely pathogenic variants. Specifically, we increasingly included each feature one by one, starting with gene-based features, then also including amino acid-based features, and finally including the nucleotide variant-based features in the same order as listed in Table 2. Each new predictor was evaluated using synthetic test sets as before. As expected, we found that the best performance was achieved by the version of VarPPUD that utilized all 38 features (accuracy of 0.813, Figure 3b). Intriguingly, although the inclusion of more features resulted in a general trend of better performance, we found that iteratively including new features individually did not always result in improved performance. For instance, including the total number of gene–chemical interactions for a gene as a feature resulted in a worse-performing predictor than including only the total number of phenotypes associated with the gene, the number of pathways the gene is a part of, and the number of unique chemicals the gene interacts with. We hypothesized that the interdependency of some features may require pairs or groups of features to be present to be exploited for a discriminatory advantage by our random forest model.

Since the version of VarPPUD that utilized all 38 features had the best performance, we next excluded each feature one-by-one to better understand the relative importance of each feature based on the overall drop in predictive performance. Specifically, we permuted the values for each feature in turn across all variants in the synthetic test sets and measured VarPPUD’s predictive accuracy (Figure 3c). We found that values for nucleotide-level CADD and VEST4 features and values for the number of total and rare diseases associated with the gene were most important for accurately determining whether a prioritized variant was likely or unlikely to be pathogenic.

### Asymmetric influence of features to model prediction

Certain values of some features may strongly implicate a variant as likely pathogenic, but certain values for an entirely different set of features may strongly indicate that a variant is unlikely pathogenic. To assess these potential differences and evaluate how and whether specific feature values impact the directionality of VarPPUD’s predictions, we next generated and analyzed Shapley Additive exPlanations (SHAP) values (Figure 3d). Briefly, for each individual prediction, a negative SHAP value for a specific feature suggests that the value of that feature contributed to a prediction of unlikely pathogenic, whereas a positive SHAP value for a specific feature suggests that that feature’s value contributed to a prediction of likely pathogenic. By overlaying SHAP values with feature values, we can see whether high or low feature values contribute to a likely or unlikely pathogenic prediction. As expected, we found that high and low CADD scores respectively corresponded to likely and unlikely pathogenic predictions. In contrast however, we found that although a small number of associated gene phenotypes often strongly contributed to a prediction of unlikely pathogenic, many associated gene phenotypes did not necessarily indicate a prediction of likely pathogenic. The number of rare and total diseases had a similarly asymmetric impact; small numbers of associated diseases with a gene resulted in a strong prediction of unlikely pathogenic, but a relatively large number of total and rare associated diseases did not indicate a prediction of likely pathogenic. It is well established that larger numbers of annotated phenotypes and associated diseases with a gene contribute to the likelihood of variants within that gene being pathogenic overall.

However, once a set of candidate variants across the genome has been prioritized for a particular patient, these features are no longer as important for selecting the exact disease-causing variant from the other candidates. Our analysis of VarPPUD’s feature importance in this manner illustrates how distinguishing likely from unlikely pathogenic variants within a set of prioritized, candidate variants is an inherently different task from identifying pathogenic variants genome-wide, and some unique features with counterintuitive values may be leveraged for this specific task. We did not find striking examples of asymmetric contributions to prediction when looking at products of feature values (Figure S1).

Finally, we assessed the mean absolute values of SHAP values per feature across all predictions as another measure of overall feature importance. We found that ranking features by importance using this metric differed from ranking features by overall accuracy drop when feature values were randomized (Figure 3e). Changes to amino acid hydrophobicity and the number of side chain interactions, for instance, had relatively high average SHAP values, yet these features did not appear to result in a large performance drop when their values were randomized. Indeed, many of the features uniquely ranked by the average SHAP value procedure appear to correspond to features where certain values can be informative for predicting one variant class, but less informative for the other variant class. Although several individual features and pairwise combinations of features contribute strongly to prediction, it is possible that some features utilized by VarPPUD were not particularly useful in distinguishing between the pathogenicity classes described here. In general, physicochemical features of amino acid substitutions influenced pathogenicity prediction to a lesser extent than gene- and nucleotide-level features.

### Patient demographics are not predictive of pathogenicity

We explored whether VarPPUD’s ability to discern likely from unlikely pathogenic variants could be improved by incorporating patient demographic information alongside the gene-, amino acid- and nucleotide-level features used (Figure 4). Although the age at first symptom onset, current age, and patient-recorded race showed promise in marginally increasing predictive capability, these values did not consistently improve model performance. Incorporating the age at first symptom onset resulted in the highest fluctuation in performance, and incorporating ethnicity and patient-recorded gender reduced overall predictive performance. Although we did not pursue further incorporation of patient-level features, it is possible that a more systematic and extensive effort to this end beyond the scope of this paper could be fruitful.

**Figure 4.**
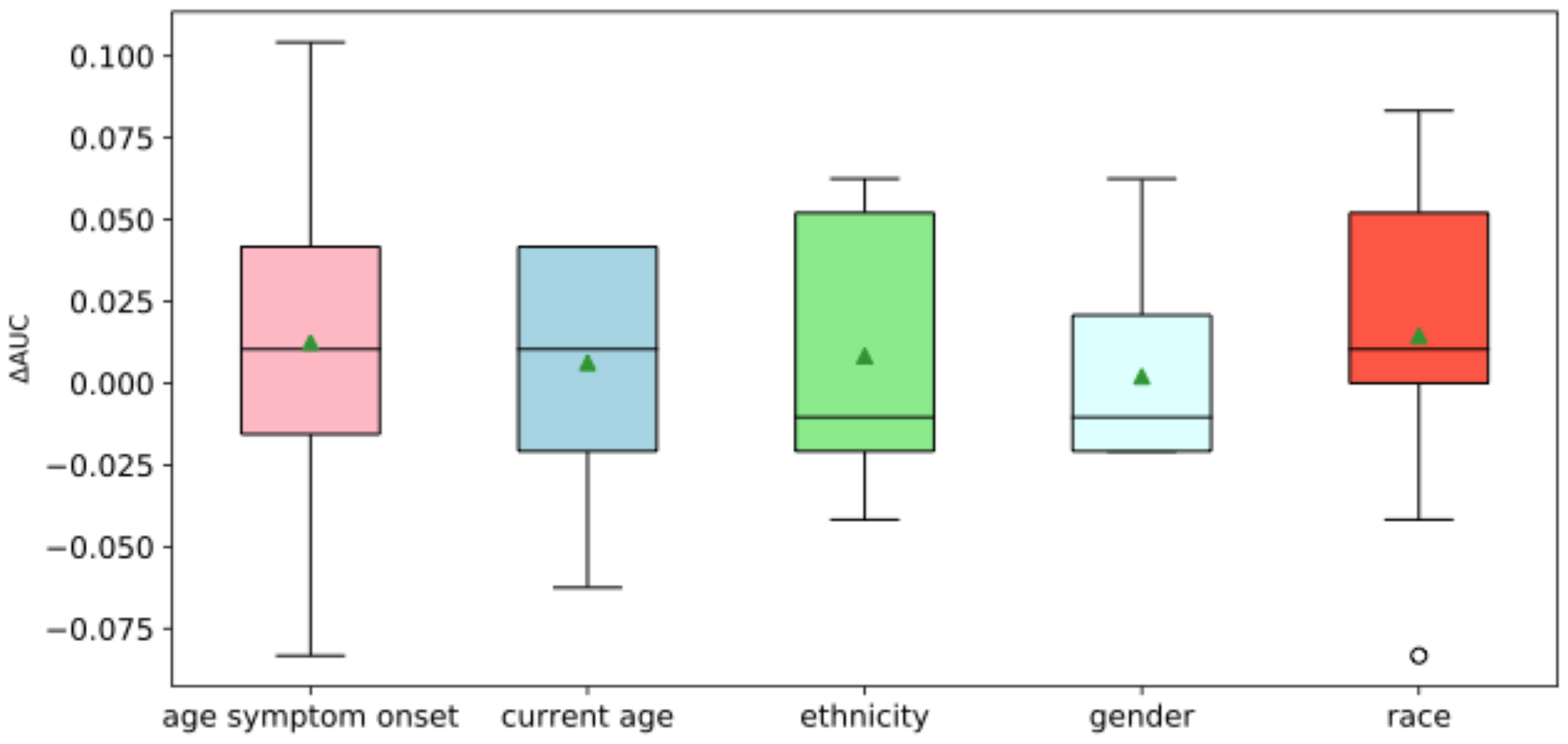
Change in performance when including patient demographics as features. We retrain VarPPUD five times, including one new demographic feature each time. We compute the area under the ROC curve (ΔAUROC) ten times for each retrained model. Horizontal lines within each boxplot indicate the median value, green triangles indicate the mean value, and white circles indicate outliers.

### Pathogenicity predictions for variants in the same genes

Candidate variants impacting the same gene across unrelated patients showing similar symptoms can sometimes be upgraded from uncertain significance to definitively causing disease. For example, three different variants in the gene *CACNA1A* (c.1360C>T, c.4055G>T, c.5018G>C) found in UDN patients within the UDN with primarily neurological symptoms were eventually confirmed as disease-causing. Similarly, three variants in *MECP2* gene (c.538C>T, c.316C>T, c.352C>T) identified in unrelated patients with symptoms akin to Rett syndrome and severe encephalopathy were reclassified as pathogenic by the overseeing clinical teams. Our tool, VarPPUD, correctly predicted these variants as likely pathogenic. However, it is important to note that not all candidate variants in the same gene are necessarily relevant to disease in every patient. For instance, two variants in the *NOD2* gene (c.2104C>T and c.2798+158C>T) were initially categorized as uncertain despite being found in two unrelated UDN patients with rheumatosis, an immune-related condition impacting the joints, muscles and ligaments. Another UDN patient with primarily immunological symptoms had a different *NOD2* variant, c.1292C>T. VarPPUD predicted all three *NOD2* variants as pathogenic. These variants have since been reclassified as likely pathogenic or pathogenic, showcasing VarPPUD’s potential as a supportive tool in reevaluating the pathogenicity of variants of uncertain significance.

## Discussion

Identifying a set of putative disease-causing variants in patients with undiagnosed genetic conditions is a challenging, multi-step process.^10^ Variant prioritization often involves the genome-wide application of *in silico* pathogenicity and deleteriousness predictors to exclude common, likely benign variants from downstream consideration. Even after filtering out likely benign variants and restricting to variants in genes that seem relevant to a patient’s phenotypic presentation, variant prioritization pipelines can still return dozens of variants of uncertain significance.^47,48^ In practice, these candidate variants are manually investigated by clinical experts and confirmed or rejected as causal through extensive functional analyses and variant matchmaking services.^49^ In our work, we accessed a unique dataset of uncertain candidate variants originally prioritized through genomic analysis of patients enrolled in the Undiagnosed Diseases Network (UDN) and eventually reclassified as benign or pathogenic. We trained a binary classifier, VarPPUD, to categorize whether any strong candidate variants were likely to be truly disease-causing or not based on nucleotide-, amino acid-, and gene-level properties. VarPPUD achieved superior performance on this classification task on both real and GAN-generated synthetic data compared to existing methods that were developed to distinguish deleterious from otherwise benign variants. Our results show that the task of selecting the single disease-causing variant from a set of already compelling variants is different from the first-pass, genome-wide prioritization task that most pathogenicity predictors have been developed for. Our model analysis revealed that some features existing utilized for a discriminatory advantage by VarPPUD, including numbers of rare or total disorders associated with specific genes, are used counter-intuitively compared to traditional predictors trained to distinguish pathogenic from benign variants genome wide. VEST, for instance, is another random forest model trained on 45,000 disease mutations from the Human Gene Mutation Database and 45,000 common, likely neutral missense variants from the Exome Sequencing Project.^50^ Despite its substantially larger training set size, VEST was less suited to distinguish likely from unlikely pathogenic variants relative to VarPPUD when applied to strong candidate variants from UDN patients.

The diagnosis rate for new patients with suspected but elusive genetic conditions hovers around 30%.^9^ Once patients have had inconclusive clinical sequencing, reanalysis or reinterpretation of their sequencing data is rarely automated, and even patients with strong candidate variants can remain in an undiagnosed limbo for many years. As the numbers of sequenced rare disease patients continues to grow, automated reanalysis and reinterpretation of variant findings will be essential for improving diagnostic rates. We propose that VarPPUD can be applied as a second-line pathogenicity predictor to select for disease-causing variants from among the sets of prioritized candidate variants in patients where progress has otherwise stalled.

We acknowledge several limitations of the work we present here. Although VarPPUD successfully makes predictions on indel variants where other missense-only predictors such as PolyPhen2 or SIFT fall short, the amino acid feature values we compute for indels may not be as biologically interpretable as those computed for SNVs. For instance, inserting an amino acid into a protein alpha-helix structure may more severely disrupt protein secondary structure than would be captured by our scoring technique. Nevertheless, we find that being able to score indel variants in this manner leads to better overall performance and enables VarPPUD to score a more realistic set of strong candidate variants that would be prioritized for a rare disease patient. Moreover, although VarPPUD makes predictions for variant types beyond SNVs, it does not score certain disease-causing genetic aberrations such as large structural rearrangements, gene duplication events, or expansions of repeat regions.

We impute feature values such as per-gene haploinsufficiency or gene essentiality for variants where those values are missing. Although imputed values are logically constrained and their inclusion improves the overall predictive performance of our model, imputed values may be incorrect and generally detract from our ability to meaningfully interpret specific predictions. Even if independent empirical data could be used to predict missing feature values instead of the MICE approach employed here, resulting values may still be misleading or clinically irrelevant.^51,52^ Independently confirming the accuracy of imputed feature values is beyond the scope of this work. Some of the most informative features used by VarPPUD are nucleotide-level deleteriousness scores produced by other variant effect predictors including CADD and PROVEAN. Meaningfully interpreting specific predictions is further hindered in cases where these ensemble scoring metrics are used to distinguish likely from unlikely pathogenic variants.

Finally, we trained VarPPUD on a relatively small set of 474 candidate variants across 413 unique genes. We show high discriminatory performance on an independent, held-out validation set and GAN-generated synthetic variant datasets, but recognize that candidate variants prioritized by other, non-UDN frameworks might be sufficiently or critically different from the UDN-based training data used here. However, the UDN is composed of 12 independent clinical research sites employing distinct, in-house prioritization pipelines, so the variants prioritized network-wide used here are likely to be generally representative of the types of variants that would be uncovered by state-of-the-art prioritization pipelines.

## Data availability

Deidentified sequencing data is regularly deposited in dbGaP (accession phs001232.v5.p2). Candidate genes and variants are submitted to MatchmakerExchange. Variant-level data, clinical significance and supporting evidence, demographic information, and phenotype information for all diagnostic variants are regularly submitted to ClinVar. Other candidate variants used to train VarPPUD are available to authorized investigators of the Undiagnosed Diseases Network.

## Code availability

The source code for testing VarPPUD’s performance on the training set, real-world validation set, and synthetic validation set can be found at https://github.com/hms-dbmi/VarPPUD.

## Ethics & Inclusion Statement

The authors declare no competing interests.

## Acknowledgements

Research reported in this manuscript was supported by the National Institutes of Health Common Fund, through the Office of Strategic Coordination/Office of the National Institutes of Health Director under Award Number U01HG007530. The content is solely the authors’ responsibility and does not necessarily represent the official views of the National Institutes of Health.

