## Supplementary for "VarPPUD: Variant post prioritization developed for undiagnosed genetic disorders"

### Supplementary Information

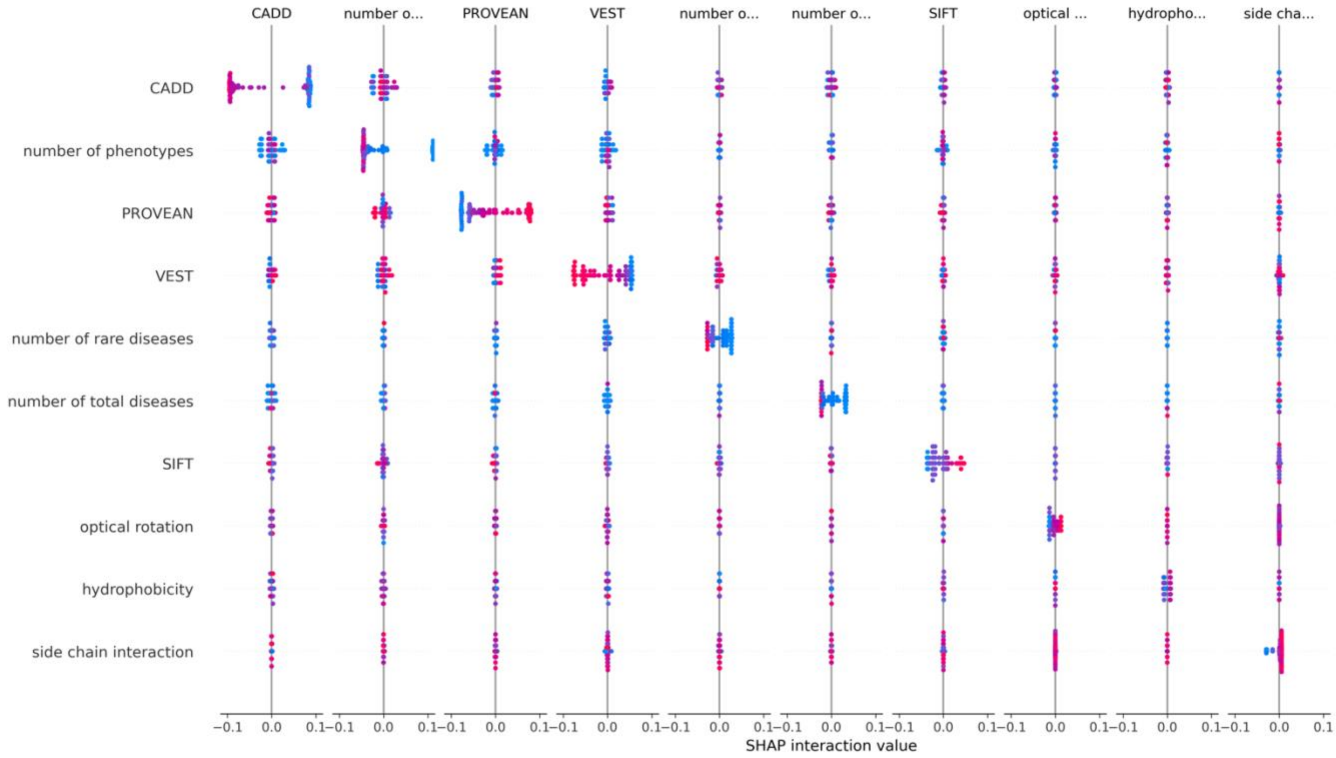


**Figure S1. SHAP interaction scores for pairs of the ten most important features predictive of variant pathogenicity in undiagnosed patients.**

**Table S1. Description of physicochemical and biochemical properties of amino acids derived from AAindex.**

| Entry | Property | Literature | Reference |
|---|---|---|---|
| RADA880108 | Polarity | Side-chain distribution coefficients between the vapor phase, cyclohexane, 1-octanol, and neutral aqueous solution | (22) |
| KLEP840101 | Net charge | Prediction of protein function from sequence properties: Discriminant analysis of a data base | (23) |
| CIDH920103 | Hydrophobicity | Hydrophobicity and structural classes in proteins | (24) |
| FAUJ880103 | Normalized van der Waals volume | Amino acid side chain parameters for correlation studies in biology and pharmacology | (25) |
| CHAM820101 | Polarizability | The structural dependence of amino acid hydrophobicity parameters | (26) |

| | | | |
|---|---|---|---|
| JOND750102 | pK-COOH | Amino acid properties and side-chain orientation in proteins: A cross correlation approach | (27) |
| FASG760104 | pK-NH2 | Handbook of Biochemistry and Molecular Biology | (28) |
| ROBB790101 | Hydration free energy | Refined models for computer simulation of protein folding: Applications to the study of conserved secondary structure and flexible hinge points during the folding of pancreatic trypsin inhibitor | (29) |
| FASG760101 | Molecular weight | Handbook of Biochemistry and Molecular Biology | (28) |
| FASG760103 | Optical rotation | Handbook of Biochemistry and Molecular Biology | (28) |
| LEVJ860101 | Secondary structure | An algorithm for secondary structure determination in proteins based on sequence similarity | (30) |
| CHAM820102 | Free energy of solution in water | The structural dependence of amino acid hydrophobicity parameters | (26) |
| FAUJ880109 | Number of hydrogen bond donors | Amino acid side chain parameters for correlation studies in biology and pharmacology | (25) |
| PONJ960101 | Average volumes of residues | Deviations from standard atomic volumes as a quality measure for protein crystal structures | (31) |
| JANJ790102 | Transfer free energy | Surface and inside volumes in globular proteins | (32) |
| WARP780101 | Average interactions per side chain atom | A survey of amino acid side-chain interactions in 21 proteins | (33) |
| KARS160101 | Number of vertices | A graph-theoretic model of single point mutations in the cystic fibrosis transmembrane conductance regulator | (34) |
| KARS160102 | Number of edges | A graph-theoretic model of single point mutations in the cystic fibrosis transmembrane conductance regulator | (34) |
| KARS160105 | Eccentricity | A graph-theoretic model of single point mutations in the cystic fibrosis transmembrane conductance regulator | (34) |
| KARS160107 | Diameter | A graph-theoretic model of single point mutations in the cystic fibrosis transmembrane conductance regulator | (34) |

| | | | |
|---|---|---|---|
| KARS160117 | Total weighted atomic number | A graph-theoretic model of single point mutations in the cystic fibrosis transmembrane conductance regulator | (34) |

**Table S2. Members of the Undiagnosed Diseases Network**

Maria T. Acosta, David R. Adams, Ben Afzali, Ali Al-Beshri, Eric Allenspach, Aimee Allworth, Raquel L. Alvarez, Justin Alvey, Ashley Andrews, Euan A. Ashley, Carlos A. Bacino, Guney Bademci, Ashok Balasubramanyam, Dustin Baldridge, Erin Baldwin, Jim Bale, Michael Bamshad, Deborah Barbouth, Pinar Bayrak-Toydemir, Anita Beck, Alan H. Beggs, Edward Behrens, Gill Bejerano, Hugo J. Bellen, Jimmy Bennett, Jonathan A. Bernstein, Gerard T. Berry, Anna Bican, Stephanie Bivona, Elizabeth Blue, John Bohnsack, Devon Bonner, Nicholas Borja, Lorenzo Botto, Lauren C. Briere, Elizabeth A. Burke, Lindsay C. Burrage, Manish J. Butte, Peter Byers, William E. Byrd, Kaitlin Callaway, John Carey, George Carvalho, Thomas Cassini, Sirisak Chanprasert, Hsiao-Tuan Chao, Ivan Chinn, Gary D. Clark, Terra R. Coakley, Laurel A. Cobban, Joy D. Cogan, Matthew Coggins, F. Sessions Cole, Brian Corner, Rosario I. Corona, William J. Craigen, Andrew B. Crouse, Vishnu Cuddapah, Precilla D'Souza, Hongzheng Dai, Kahlen Darr, Surendra Dasari, Joie Davis, Margaret Delgado, Esteban C. Dell'Angelica, Katrina Dipple, Daniel Doherty, Naghmeh Dorrani, Jessica Douglas, Emilie D. Douine, Dawn Earl, Lisa T. Emrick, Christine M. Eng, Cecilia Esteves, Kimberly Ezell, Elizabeth L. Fieg, Paul G. Fisher, Brent L. Fogel, Jiayu Fu, William A. Gahl, Rebecca Ganetzky, Emily Glanton, Ian Glass, Page C. Goddard, Joanna M. Gonzalez, Andrea Gropman, Meghan C. Halley, Rizwan Hamid, Neal Hanchard, Kelly Hassey, Nichole Hayes, Frances High, Anne Hing, Fuki M. Hisama, Ingrid A. Holm, Jason Hom, Martha Horike-Pyne, Alden Huang, Yan Huang, Anna Hurst, Wendy Introne, Gail P. Jarvik, Suman Jayadev, Orpa Jean-Marie, Vaidehi Jobanputra, Oguz Kanca, Yigit Karasozen, Shamika Ketkar, Dana Kiley, Gonench Kilich, Eric Klee, Shilpa N. Kobren, Isaac S. Kohane, Jennefer N. Kohler, Bruce Korf, Susan Korrick, Deborah Krakow, Elijah Kravets, Seema R. Lalani, Christina Lam, Brendan C. Lanpher, Ian R. Lanza, Kumarie Latchman, Kimberly LeBlanc, Brendan H. Lee, Kathleen A. Leppig, Richard A. Lewis, Pengfei Liu, Nicola Longo, Joseph Loscalzo, Richard L. Maas, Ellen F. Macnamara, Calum A. MacRae, Valerie V. Maduro, AudreyStephannie Maghiro, Rachel Mahoney, May Christine V. Malicdan, Rong Mao, Ronit Marom, Gabor Marth, Beth A. Martin, Martin G. Martin, Julian A. Martínez-Agosto, Shruti Marwaha, Allyn McConkie-Rosell, Ashley McMinn, Matthew Might, Mohamad Mikati, Danny Miller, Ghayda Mirzaa, Breanna Mitchell, Paolo Moretti, Marie Morimoto, John J. Mulvihill, Lindsay Mulvihill, Mariko Nakano-Okuno, Stanley F. Nelson, Serena Neumann, Dargie Nitsuh, Donna Novacic, Devin Oglesbee, James P. Orengo, Laura Pace, Stephen Pak, J. Carl Pallais, Neil H. Parker, LéShon Peart, Leoyklang Petcharet, John A. Phillips III, Filippo Pinto e Vairo, Jennifer E. Posey, Lorraine Potocki, Barbara N. Pusey Swerdzewski, Aaron Quinlan, Daniel J. Rader, Ramakrishnan Rajagopalan, Deepak A. Rao, Anna Raper, Wendy Raskind, Adriana Rebelo, Chloe M. Reuter, Lynette Rives, Lance H. Rodan, Martin Rodriguez, Jill A. Rosenfeld, Elizabeth Rosenthal, Francis Rossignol, Maura Ruzhnikov, Marla Sabaii, Jacinda B. Sampson, Timothy Schedl, Lisa Schimmenti, Kelly Schoch, Daryl A. Scott, Elaine Seto, Vandana Shashi, Emily Shelkowitz, Sam Sheppeard, Jimann Shin, Edwin K. Silverman, Giorgio Sirugo, Kathy Sisco, Tammi Skelton, Cara Skraban, Carson A. Smith, Kevin S. Smith, Lilianna Solnica-Krezel, Ben Solomon, Rebecca C. Spillmann, Andrew Stergachis, Joan M. Stoler, Kathleen Sullivan, Shamil R. Sunyaev, Shirley Sutton, David A. Sweetser, Virginia Sybert, Holly K. Tabor, Queenie Tan, Arjun

Tarakad, Herman Taylor, Mustafa Tekin, Willa Thorson, Cynthia J. Tifft, Camilo Toro, Alyssa A. Tran, Rachel A. Ungar, Adeline Vanderver, Matt Velinder, Dave Viskochil, Tiphanie P. Vogel, Colleen E. Wahl, Melissa Walker, Nicole M. Walley, Jennifer Wambach, Michael F. Wangler, Patricia A. Ward, Daniel Wegner, Monika Weisz Hubshman, Mark Wener, Tara Wenger, Monte Westerfield, Matthew T. Wheeler, Jordan Whitlock, Lynne A. Wolfe, Heidi Wood, Kim Worley, Shinya Yamamoto, Zhe Zhang, Stephan Zuchner

### Supplementary Information References

14. Ng P C, Henikoff S. SIFT: Predicting amino acid changes that affect protein function[J]. Nucleic acids research, 2003, 31(13): 3812-3814.
15. Rentzsch P, Witten D, Cooper G M, et al. CADD: predicting the deleteriousness of variants throughout the human genome[J]. Nucleic acids research, 2019, 47(D1): D886-D894.
16. Graham J W. Missing data analysis: Making it work in the real world[J]. Annual review of psychology, 2009, 60: 549-576.
17. Azur M J, Stuart E A, Frangakis C, et al. Multiple imputation by chained equations: what is it and how does it work?[J]. International journal of methods in psychiatric research, 2011, 20(1): 40-49.
18. Breiman L. Random forests[J]. Machine learning, 2001, 45(1): 5-32.
19. Xu L, Skoularidou M, Cuesta-Infante A, et al. Modeling Tabular data using Conditional GAN[J]. Advances in Neural Information Processing Systems, 2019, 32: 7335-7345.
20. Winter E. The shapley value[J]. Handbook of game theory with economic applications, 2002, 3: 2025-2054.
21. Lundberg S M, Lee S I. A Unified Approach to Interpreting Model Predictions[J]. Advances in Neural Information Processing Systems, 2017, 30: 4765-4774.
22. Radzicka A, Wolfenden R. Comparing the polarities of the amino acids: side-chain distribution coefficients between the vapor phase, cyclohexane, 1-octanol, and neutral aqueous solution[J]. Biochemistry, 1988, 27(5): 1664-1670.
23. Klein P, Kanehisa M, DeLisi C. Prediction of protein function from sequence properties: Discriminant analysis of a data base[J]. Biochimica et Biophysica Acta (BBA)-Protein Structure and Molecular Enzymology, 1984, 787(3): 221-226.
24. Cid H, Bunster M, Canales M, et al. Hydrophobicity and structural classes in proteins[J]. Protein Engineering, Design and Selection, 1992, 5(5): 373-375.
25. FAUCHÈRE J L U C, Charton M, Kier L B, et al. Amino acid side chain parameters for correlation studies in biology and pharmacology[J]. International journal of peptide and protein research, 1988, 32(4): 269-278.
26. Charton M, Charton B I. The structural dependence of amino acid hydrophobicity parameters[J]. Journal of theoretical biology, 1982, 99(4): 629-644.
27. Jones D D. Amino acid properties and side-chain orientation in proteins: a cross correlation approach[J]. Journal of theoretical biology, 1975, 50(1): 167-183.
28. Fasman G D. Practical handbook of biochemistry and molecular biology[M]. CRC press, 1989.
29. Robson B, Osguthorpe D J. Refined models for computer simulation of protein folding: Applications to the study of conserved secondary structure and flexible hinge points during the folding of pancreatic trypsin inhibitor[J]. Journal of molecular biology, 1979, 132(1): 19-51.
30. Levin J M, Robson B, Garnier J. An algorithm for secondary structure determination in proteins based on sequence similarity[J]. FEBS letters, 1986, 205(2): 303-308.
31. Pontius J, Richelle J, Wodak S J. Deviations from standard atomic volumes as a quality measure for protein crystal structures[J]. Journal of molecular biology, 1996, 264(1): 121-136.
32. Janin J. Surface and inside volumes in globular proteins[J]. Nature, 1979, 277(5696): 491-492.
33. Warme P K, Morgan R S. A survey of amino acid side-chain interactions in 21 proteins[J]. Journal of molecular biology, 1978, 118(3): 289-304.

34. Kakraba S, Knisley D. A graph-theoretic model of single point mutations in the cystic fibrosis transmembrane conductance regulator[J]. Journal of Advances in Biotechnology, 2016, 6(1): 780-786.
